# Early identification of postpartum depression using demographic, clinical, and digital phenotyping

**DOI:** 10.1101/2020.06.26.20140673

**Authors:** Lisa Hahn, Simon B. Eickhoff, Ute Habel, Elmar Stickeler, Patricia Schnakenberg, Tamme W. Goecke, Susanne Stickel, Matthias Franz, Juergen Dukart, Natalia Chechko

## Abstract

**Background:** Postpartum depression (PPD) and adjustment disorder (AD) affect up to 25% of women after childbirth. However, there are no accurate screening tools for either disorder to identify at-risk mothers and enable them to benefit from early intervention.

**Aims:** To evaluate combinations of anamnestic, clinical and remote assessments for an early and accurate identification of PPD and AD.

**Method:** Two cohorts of mothers giving birth were included in the study (N=308 and N=193). At baseline, participants underwent a detailed anamnestic and clinical interview. Remote assessments were collected over twelve weeks comprising mood and stress levels as well as depression and attachment scores. At twelve weeks postpartum, an experienced clinician assigned the participants to three distinct groups: women with PPD, women with AD, and healthy controls (HC). Combinations of these assessments were assessed for an early an accurate detection of PPD and AD in the first cohort and, after pre-registration, prospectively validated in the second cohort.

**Results:** Combinations of postnatal depression, attachment (for AD) and mood scores at week 3 achieved balanced accuracies of 93% and 79% for differentiation of PPD and AD from HC in the prospective validation cohort. Differentiation between AD and PPD, with a balanced accuracy of 73 % was possible at week 6 based on mood levels only.

**Conclusions:** Combinations of in clinic and remote self-assessments allowed for early and accurate detection of PPD and AD as early as three weeks postpartum, enabling early intervention to the benefit of both mothers and children.

## Introduction

The postpartum period poses the highest risk for women to develop a mental disorder (1), with postpartum depression (PPD) being the most frequent one (2). PPD is defined as a major depressive disorder (MDD) occurring in direct relation (within 4 weeks postpartum) to childbirth in the DSM-5 (3). Early diagnosis and treatment of PPD can substantially improve the outcome, prevent relapse, and minimize the associated emotional and financial burden (4). Maternal mental health is a reliable predictor of child’s cognitive development and subsequent achievements (5). The risk of a mother-to-child transmission of the vulnerability to depression (6,7), through genetic as well as other factors such as depression-related effects on parenting (8), is particularly high. Successful treatment of maternal depression alleviates the risk of childhood behavioral problems (9).

### Diagnostic difficulties

PPD is often overlooked during postnatal visits, missing the critical window for early intervention (10,11). One reason is that low mood in the early postpartum period is largely deemed “normal” with 50 to 80% of new mothers experiencing initial sadness (i.e. postpartum blues), primarily due to dramatically plunging hormone levels at parturition (12). Adjustment disorder (AD) in reaction to postpartum stress is another postpartum condition with similar symptoms. The crucial difference to PPD is that the severity of AD does not meet the criteria for depression at any time point. In the clinical context, AD needs to be considered as an important differential diagnosis to PPD (13).

### Risk factors and previous studies

History of mental illness, vulnerability to hormonal changes, psychological and social distress, baby blues, premenstrual syndrome (PMS), unwanted pregnancy, traumatic birth experience and stressful life events are all associated with an increased risk of PPD (11,12,14). It is of crucial importance to evaluate the relative and combined predictive value of these factors for development of PPD. Previous studies aiming at prediction of PPD focused either on time points in the late postpartum period (e.g. after 8 to 32 weeks) (15) or only on single time points, thereby ignoring symptom dynamics or convolving PPD with major depression or AD (16). Detailed in-clinic assessments are costly and burdensome, providing the likely reason for the cross-sectional nature of most previous studies. Online remote self-assessments may provide an easy means of obtaining the relevant information on symptom dynamics in individual patients.

### The current study

Here, we recruited two cohorts of mothers giving birth and followed them longitudinally over twelve weeks to explore whether an accurate prediction of PPD is feasible based on socio-demographic and clinical-anamnestic information as well as early symptom dynamics using remote mood and stress assessments. Data from the first cohort were used to identify combinations of demographic and clinical data achieving highest accuracy for early identification and differentiation of PPD and AD using a machine learning approach. In this cohort, we identified and trained the optimal model for individual diagnostic prediction. The model and approach were pre-registered and evaluated against an independent validation cohort to obtain unbiased performance estimates of the proposed algorithm.

## Methods

[Table 1]

**Table 1.** Demographic and anamnestic data for the first and second cohort.

| <u>Anamnestic variable</u> | <u>First cohort</u> |  |  |  | <u>Second cohort</u> |  |  |  |
| --- | --- | --- | --- | --- | --- | --- | --- | --- |
|  | <u>HC</u> | <u>PPD</u> | <u>AD</u> | <u>Statistical test</u> | <u>HC</u> | <u>PPD</u> | <u>AD</u> | <u>Statistical test</u> |
| Age (in years) | 31.9 ± 4.61<br>N = 247 | 30.4 ± 5.51<br>N = 28 | 31.4 ± 5.09<br>N = 33 | $X^2(2, N= 308) = 2.60$<br>$p = .27$ | 33.1 ± 4.31<br>N = 146 | 31.8 ± 7.04<br>N = 16 | 31.5 ± 5.42<br>N = 30 | $X^2(2, N= 193) = 3.40$<br>$p = .18$ |
| Education (in years) | 13.8 ± 2.89<br>N = 240 | 12.4 ± 2.85<br>N = 27 | 13.4 ± 4.67<br>N = 33 | $X^2(2, N= 300) = 3.04$<br>$p = .22$ | 14.6 ± 3.22<br>N = 139 | 14.3 ± 2.96<br>N = 16 | 14.08 ± 2.56<br>N = 26 | $X^2(2, N= 179) = 0.77$<br>$p = .68$ |
| Personal psychiatric history (no/yes) | 220/27 | 16/12 | 19/14 | $X^2(2, N= 308) = 34.5$<br>$p < .001$ <sup>*1,2</sup> | 118/26 ■ | 6/10 | 15/15 | $X^2(2, N= 190) = 24.2$<br>$P < .001$ <sup>*1,2</sup> |
| Familial psychiatric history (no/yes) | 194/53 | 16/12 | 18/15 | $X^2(2, N= 308) = 13.3$<br>$p = .001$ <sup>*1,2</sup> | 112/33 | 8/8 | 65/14 | $X^2(2, N= 193) = 10.8$<br>$p = .005$ <sup>*2</sup> |
| Birth complications (no/yes) | 209/37 | 20/8 | 24/9 | $X^2(2, N= 307) = 5.57$<br>$p = .062$ | 121/26 | 10/6 | 25/5 | $X^2(2, N= 193) = 3.80$<br>$p = .15$ |
| Subjective birth-related psychological traumas (no/yes) | 215/29 | 19/9 | 20/13 | $X^2(2, N= 305) = 21.1$<br>$p < .001$ <sup>*1,2</sup> | 124/14 | 12/4 | 23/7 | ° |
| PMS (no PMS/mild PMS/PMS) | 111/84/29 | 4/12/12 | 7/16/10 | $X^2(4, N= 285) = 27.9$<br>$p < .001$ <sup>*1</sup> | 83/44/14 | 3/4/8 | 9/14/6 | $X^2(2, N= 185) = 26.6$<br>$p < .001$ <sup>*°</sup> |
| Postpartum blues (no/yes) | 151/93 | 8/20 | 7/26 | $X^2(2, N= 305) = 27.7$<br>$p < .001$ <sup>*1,2</sup> | 102/45 | 0/16 ■ | 9/21 | $X^2(2, N= 193) = 39.4$<br>$p < .001$ <sup>*°</sup> |
| Stressful life events<br>(number)<br>(no/yes) | 0.81 ± 1.27 | 1.46 ± 1.71 | 1.19 ± 1.18 |  | 1 ± 1.35 | 2.44 ± 2.06 | 1.97 ± 1.88 |  |
| | 144/103 | 11/17 | 12/20 | $X^2(2, N= 307) = 7.78$<br>$p = .020$ | 71/74 | 2/14 | 8/22 | $X^2(2, N= 191) = 11.5$<br>$p = .003$ <sup>*1</sup> |
| Breastfeeding T4 (no/yes) | 63/182 | 14/14 | 8/25 | $X^2(2, N= 306) = 7.62$<br>$p = .022$ <sup>*1</sup> | 41/96 | 9/7 | 9/20 | $X^2(2, N= 182) = 4.56$<br>$p = .10$ |
| Hamilton Depression Rating Scale T4 | -- | 13.2 ± 2.88<br>N = 27 | -- |  | -- | 14.6 ± 4.18<br>N = 16 | -- | -- |

° No statistical analysis possible due to low expected cell counts.
\* Bonferroni-corrected significant difference ( $p < .05$ ) between <sup>1</sup> HC and PPD, <sup>2</sup> between HC and AD and/or <sup>3</sup> between PPD and AD
■ Significant group difference ( $p < .05$ ) between first and validation cohort
**AD – adjustment disorder, HC – healthy controls, PMS – premenstrual syndrome, PPD – postpartum depression**

### First cohort and study design

To identify the best predictors of PPD, a first cohort of 308 mothers (mean age = 31.7 ± 4.76) was recruited following childbirth at the University Hospital Aachen. The main exclusion criteria were a depressive episode during pregnancy and specific child health conditions (for details see Supplementary material). The recruitment was conducted at the Department of Gynecology and Obstetrics within the first two to five days postpartum. Written informed consent was obtained from all subjects/patients. The authors assert that all procedures contributing to this work comply with the ethical standards of the relevant national and institutional committees on human experimentation and with the Helsinki Declaration of 1975, as revised in 2008. All procedures involving human subjects/patients were approved by the Institutional Review Board of the Medical Faculty of RWTH Aachen University (EK 208/15). The study design comprised follow-up for 12 weeks with evaluation at five time points each three weeks apart (T0 - T4) (Figure S1). Evaluations were conducted at the clinic for T0 and T4 and via remote online questionnaires for T1 to T3. All women were asked to complete mood and stress assessments (scale from one to ten, ten being high) online on a bi-daily basis.

A clinical interview was conducted at T0 to ascertain current conditions. At T4, an experienced psychiatrist conducted a second clinical interview for a final diagnosis. Based on this interview, participants were assigned into one of three groups: healthy controls (HC, N = 247, 80.2%) without any sign of depression during the whole observation period, and women meeting DSM-5 criteria for PPD (N = 28, 9.1%) or AD (N = 33, 10.7%) (3). In case of a depression, the Hamilton Depression Rating Scale (17) was administered. Clinical interviews were based on the DSM-5 (3).

An anamnestic questionnaire was used to obtain additional information about personal and socioeconomic status, psychiatric history, current pregnancy, child, breastfeeding at T0, postpartum blues (T4), premenstrual syndrome (PMS) (18) (T4), subjective quality of support at home (T4), and breastfeeding at T4 (Table 1, Table S1). The Stressful Life Events Screening Questionnaire (SLESQ) (19) was collected to assess encounter with stressful life events (T0) (Table 1). The Edinburgh Postnatal Depression Scale (EPDS) (20) was collected at all time points (T0-T4). Maternal attachment was evaluated from T1 through T4 using the Maternal Postnatal Attachment Scale (MPAS) (21).

### Second cohort

For the second cohort, further referred to as validation cohort, 193 mothers (mean age = 32.7 ± 4.78) were recruited following the same protocol and study design as for the first cohort (Figure S1). The prevalence rates in the validation cohort were 76.2% for HC (N = 147), 8.29% for PPD (N = 16), and 15.5% for AD (N = 30).

### Univariate analyses of the first cohort

All data were analyzed using MATLAB R2018a, Python Jupyter Notebook 5.6.0, IBM SPSS Statistics 22 and jamovi 1.0.5.0 (22). Chi-square tests were performed to compare categorical variables across the groups in the first cohort. For continuous variables, logistic regressions were computed. Weekly mood and stress levels were calculated by averaging the corresponding bi-daily assessments. Mood-stress difference scores were calculated as the difference between both z-transformed variables to estimate individual discrepancies between perceived stress and mood (i.e. z-score mood minus z-score stress). Changes from baseline and the preceding week were computed for these variables. Dynamic changes in mood, stress, mood-stress difference, MPAS, and EPDS were analyzed using mixed effects repeated-measures analyses of variance with week as within-subject and group as between-subject variable including an interaction term. Bonferroni-corrected independent-samples t-tests were carried out for post-hoc pair-wise group comparisons at different time points. Receiver operating characteristic (ROC) curves and their associated area under the curve (AUC) (within-sample) for differentiation between the three groups were computed for each measure per week.

### Identification of most predictive combinations in the first cohort

Next, we aimed to evaluate if and which combinations of demographic and clinical-anamnestic factors, mood, stress, MPAS and EPDS allow for an accurate differentiation between HC, PPD and AD in the first cohort. To that end, we used a logistic regression classifier (MATLAB built-in mnrfit and mnrval functions, no parameter optimization needed) performing 1000 repetitions of strict three-fold cross-validation. The classification was performed for each pair-wise group comparison separately. Low-variance variables (i.e. family status, breastfeeding T0, education, completed professional education, income, and psychiatric diagnosis in previous pregnancy) were excluded from the analysis in the whole sample. Independent samples t-tests were performed in the training data to select the baseline variables to be included in the classifier (p<.05).

To identify the most sensitive combinations for early identification of PPD, the following nine feature combinations were evaluated: [1] baseline anamnestic data alone, [2] mood scores, [3] stress scores, [4] mood-stress difference scores, [5] mood scores incl. changes (change to baseline and to preceding week), [6] stress scores incl. change scores, [7] mood-stress difference scores incl. changes, [8] combination of mood and stress scores incl. changes, [9] and combination of mood, stress, and mood-stress difference scores incl. changes. Combinations [1] to [9] were evaluated either alone or in combination with EPDS scores, MPAS scores or both. Additionally, all combinations with features [2] to [9] were evaluated with and without inclusion of baseline anamnestic information.

Balanced accuracies, sensitivities, specificities, positive and negative predictive values as well as receiver operating characteristic (ROC) curves including the area under the curve (AUC) were computed. The best performing combination (high balanced accuracy at earliest possible time-point) for each pair-wise comparison was selected for replication analysis. A logistic regression was computed for the selected combination using all participants. These results of the first cohort along with the validation plan were pre-registered on https://osf.io/ecmrp?view_only=6feb8e89818445a0b675621c8f22ba82. The obtained coefficients were applied to the prospectively collected validation cohort.

### Application to the validation cohort

The selected and preregistered model as trained on the first dataset was then used to predict diagnoses in the independent validation cohort (Table S6). The class probability *p* for the validation cohort was obtained using the following standard logistic regression formula, where β denotes the coefficients and *X* the included features:

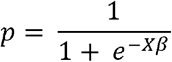

As for the validation cohort, we computed balanced accuracy, sensitivity, specificity, AUC, ROC, and positive and negative predictive value by comparing predicted versus actual group labels. To obtain a chance level spread estimate for the classifier, we randomly permuted the ‘predicted’ labels 1000 times across the validation cohort recomputing all performance measures and their 95% confidence interval.

## Results

### Anamnestic and baseline group comparisons

In the first cohort, PPD and AD were associated with personal (p < .001 for HC vs. PPD and HC vs. AD) and familial psychiatric history (p = .036 for HC vs. PPD, p = .009 for HC vs. AD), subjective birth-related psychological traumas (p = .024 for HC vs. PPD, p < .001 for HC vs. AD), and postpartum blues (p = .003 for HC vs. PPD, p < .001 for HC vs. AD) (Table 1, S1 and S2). A higher PMS prevalence (p = .012 for HC vs. PPD) and reduced breastfeeding at T4 were observed in PPD compared to HC (p = .021). No differences were seen between PPD and AD. Similar effects were observed in the validation cohort for all anamnestic factors (Table 1, Table S1).

### Univariate analyses of the first cohort

Both PPD and AD showed a distinct pattern in weekly mood, stress, and mood-stress difference scores over the course of 12 weeks (significant time by diagnosis interactions – mood: F(13.8,1303) = 16.3, p < .001; stress: F(11.3,1026) = 9.85, p < .001; mood-stress difference: F(13.1,1162) = 17.3, p < .001) (Figure 1A-C). The groups differed significantly in mood and mood-stress difference at all weeks (p = .004 for mood-stress baseline, all other p < .001) (see Tables S3-5). For stress, the difference was significant at all weeks except for baseline (all p < .001).

**Figure 1.**
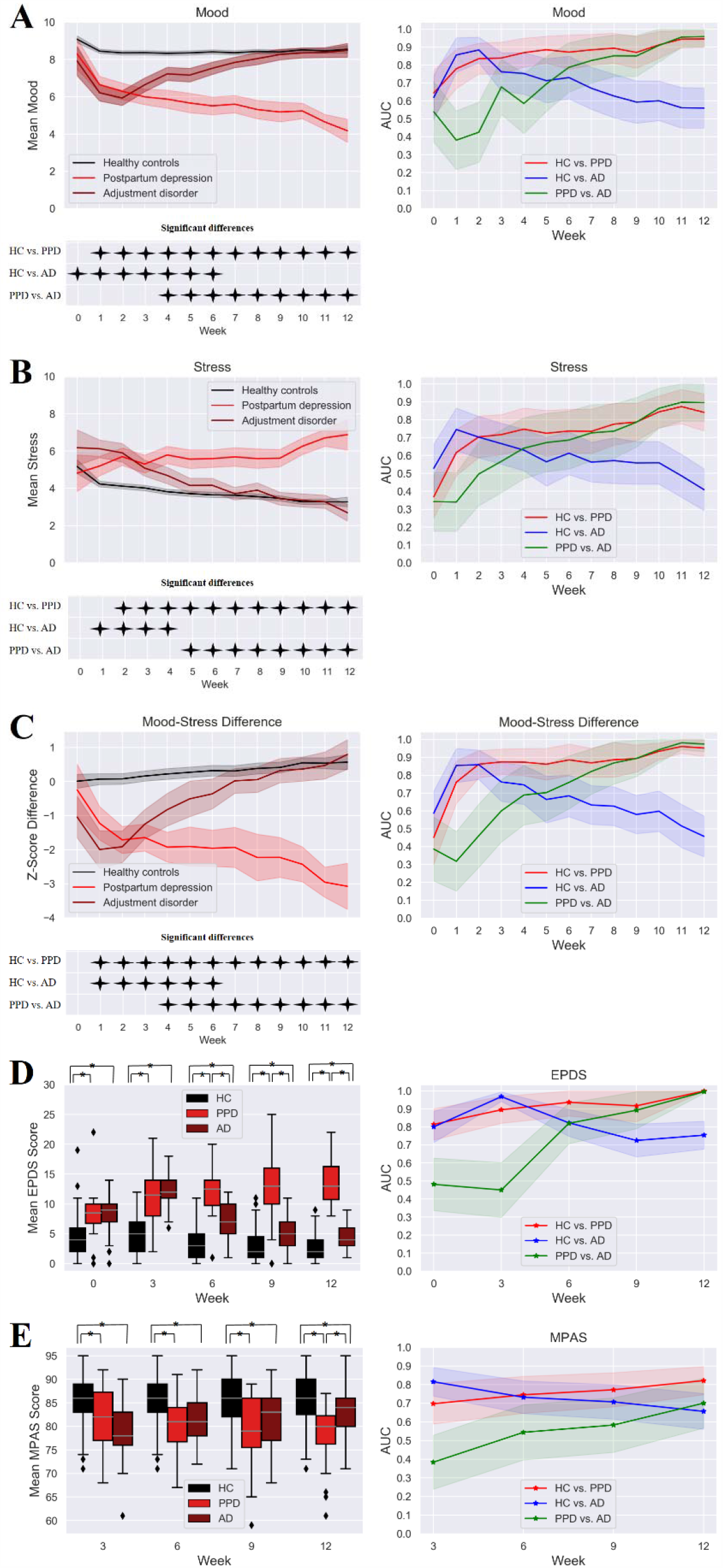
Mood, stress, mood-stress difference, EPDS, and MPAS scores. Weekly mood (A), stress, (B) and mood-stress difference scores (C) incl. 95% confidence intervals, results of the simple effects analyses, and within-sample AUCs incl. 95% confidence interval for each group comparison. EPDS (D) and MPAS (E) mean scores and associated within-sample AUCs for each time point and group separately incl. their standard error and 95% confidence interval. Statistically significant t-tests for group comparisons are marked with *.

PPD had significantly lower mood levels compared to HC at all weeks except for baseline (Figure 1A). AD had significantly lower mood relative to HC from baseline until week 6 reaching the highest difference at week 2. PPD had lower mood compared to AD from week 4 through week 12. Stress levels were significantly higher in PPD compared to HC from week 2 through week 12 and compared to AD between week 5 and week 12. AD had higher stress levels relative to HC from week 1 until week 4 (Figure 1B). Mood-stress difference was significantly different between HC and PPD from week 1 through week 12, between HC and AD from week 1 until week 6, and between PPD and AD from week 4 through week 12 (Figure 1C).

Both EPDS and MPAS showed significant time by diagnosis interactions (EPDS: F(6.87,1034) = 34.4, p < .001; MPAS: F(5.35,805) = 8.24, p < .001) with a significant between-group difference at all weeks (all p<.001) (Figure 1D-E). EPDS scores were significantly lower in HC compared to PPD and AD at all time-points (T0 - T4) (p < .001). The difference between PPD and AD was significant from T2 until T4 with higher EPDS scores in PPD women (p < .001). MPAS scores were significantly lower at all time points (T1 - T4) in PPD (p < .001) and AD (p < .001 for T1-T3, p = .008 for T4) compared to HC. Lower MPAS scores were observed in PPD compared to AD at T4 (p = .001).

### Prediction in the first cohort

Next, we evaluated when and which combinations of anamnestic, mood, stress, EPDS, and MPAS data allow for reliable differentiation between PPD, AD and HC. The outcomes of all evaluated combinations are summarized in Tables S7-14. For differentiation of PPD from HC, a high balanced accuracy of 87% was achieved at week 3 using a combination of baseline EPDS and follow-up EPDS and mood levels at week 3 (Table 2, Figure 2A and Table S7). Best early differentiation between AD and HC with a 91% balanced accuracy was also achieved at week 3 using a combination of baseline EPDS and EPDS, MPAS and mood scores at week 3 (Table 2, Figure 2B, and Table S8). A reasonable differentiation of AD and PPD with a balanced accuracy of 76% was only achieved at week 6 using only the mood levels (Table 2, Figure 2C and Table S9). Logistic regression coefficients were trained with these combinations using the first cohort and applied to predict the diagnostic labels in the validation cohort (Table S6).

**Table 2.**
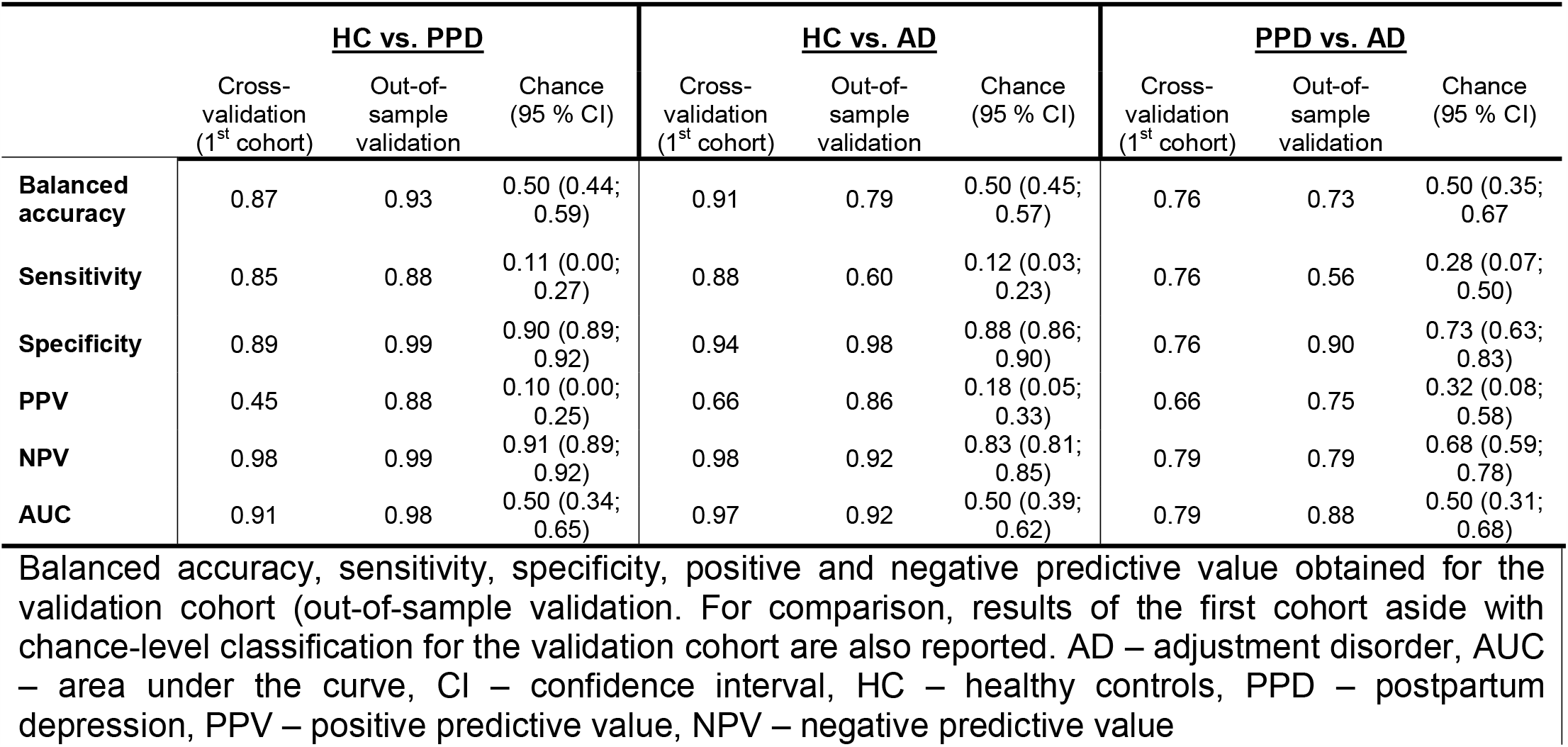
Results of prediction for the first and validation cohort

**Figure 2.**
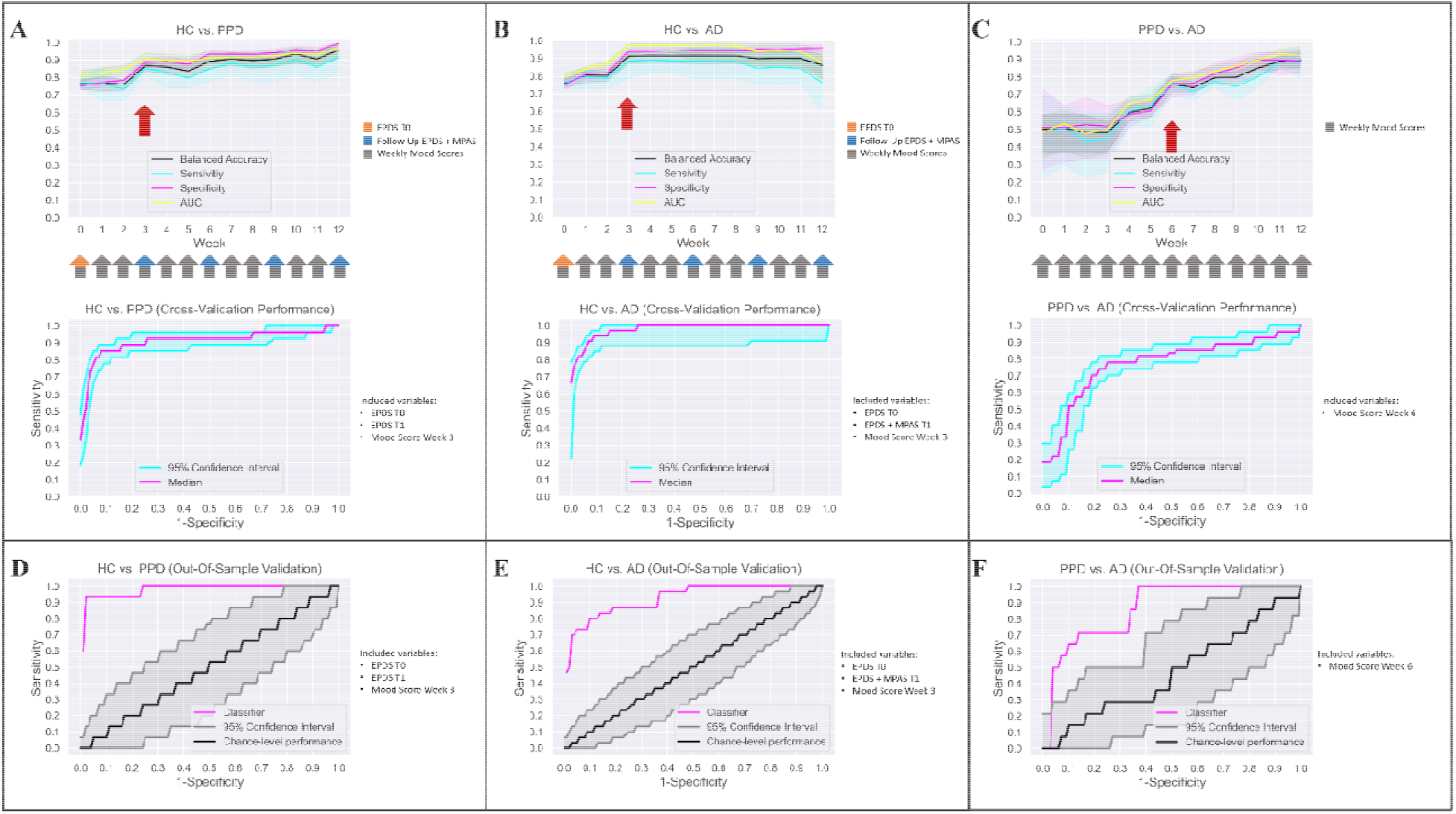
Results of machine learning analysis. Balanced accuracy, sensitivity, specificity and out-of-sample AUC for each group comparison are displayed for the first cohort (A-C). For HC vs. PPD (A), the values are displayed for EPDS at baseline and follow-up incl. mood scores. For HC vs. AD (B), the values are displayed for EPDS at baseline, EPDS and MPAS at follow-up incl. mood scores. For PPD vs. AD (C), the values are displayed for mood scores. (D-F) AUCs obtained for the validation cohort are displayed for the classifier selected based on results from the first cohort aside with chance-level performance.

### Prediction in the validation cohort

The classifier trained on the first cohort for differentiation of HC and PPD reached a high balanced accuracy of 93% in the validation cohort with a sensitivity of 88% and specificity of 99% (Table 2, Figure 2D). The classifier differentiating HC and AD reached a balanced accuracy of 79% with a high specificity (98%) but only moderate sensitivity (60%) (Table 2, Figure 2E).

For PPD and AD differentiation, the selected classifier reached a balanced accuracy of 73%, again with high specificity (90%) but only low sensitivity (56%) (Table 2, Figure 2F).

## Discussion

Here, we adopted a within- and out-of-sample validation study design to identify combinations of demographic and clinical factors allowing for early and accurate identification and differentiation of PPD and AD in two large cohorts of postpartum women. In both cohorts high accuracy is achieved at week 3 for identification of PPD and AD compared to HC using a simple combination of EPDS, mood and MPAS (for AD) assessments. In contrast, differentiation of PPD and AD is possible only from week 6 based solely on mood levels.

In both cohorts, the prevalence of PPD was slightly lower than the 10-20 % reported in the literature (23,24). As the focus of our study was on prediction of PPD, we purposely excluded women with depression during pregnancy, which may explain the lower prevalence. Furthermore, studies estimating early prevalence of PPD may have included women with AD. In line with previous research, we found postpartum blues, psychiatric history, subjective birth-related psychological traumas, and premenstrual syndrome to be significant risk factors for PPD (14,25,26).

Interestingly, no differences between the PPD and AD groups were found with respect to risk factors, suggesting that similar mechanisms may be involved in the generation of initial depressive symptoms in both groups. Over the observation period, stress levels continuously increased in women with PPD whilst they normalized after about five weeks in AD. Descriptively, mood levels in AD followed the stress levels normalizing only after about seven weeks. The temporal delay is in line with the interpretation that reductions in stress may contribute to the recovery observed in mood. The increase in stress levels and the simultaneous decline in mood levels in PPD may indicate the contribution of stress-mediated components in line with previous studies reporting parenting stress among the most important postpartum factors (27,28). Whilst not a causal factor on its own, parenting stress is likely to increase vulnerability to depression in high-risk individuals.

Similarly, PPD and AD displayed distinct temporal courses of EPDS and attachment scores as measured by MPAS. The EPDS temporal dynamics were highly similar to the observed stress and mood levels. The initially lowest attachment scores were found to increase in AD while PPD maintained the low attachment levels throughout the study. These observations underscore the necessity of longitudinal monitoring of both measures to better characterize the dynamic relationship between depressed mood and maternal attachment (29,30). Differences in MPAS and EPDS remained significant between AD and HC at all time points. According to recent findings, child neurodevelopment is affected by maternal depressive symptoms even when they do not exceed clinical thresholds (31,32). Our observations emphasize the need for further detailed evaluation of potential consequences also for the AD group.

A combination of baseline EPDS and week 3 remote follow-up EPDS and mood scores achieved about 90% balanced accuracy for early identification of PPD as compared to HC. The same combination with addition of MPAS achieved a similar accuracy for early identification of AD. Both findings were largely confirmed in the validation cohort with an accuracy reduction from 90 to 80% seen only for differentiation of AD and HC. None of the evaluated combination allowed for an accurate early differentiation of PPD and AD with all classifiers performing close to chance level until week 5. A reasonable differentiation of both groups was only achieved through mood scores at week 6 with a moderately high accuracy but a high specificity for PPD as confirmed in the validation cohort. Our classification results suggest that a simple stepwise procedure including remote mood, EPDS and MPAS assessments may be a promising approach towards early identification of PPD. Whilst week 3 remote testing provides a high accuracy and a particularly high specificity for detection of both populations at risk, week 6 data additionally allow for further differentiation between PPD and AD.

In summary, by means of a longitudinal approach we identify and validate combinations of remote assessments allowing for early and accurate identification and differentiation of PPD and AD using a step-wise procedure. Behavioral and clinical time courses over 12 weeks provided important insight into the development and interaction of mood, stress and maternal sensitivity in the first weeks postpartum.

## Data Availability

The data that support the findings of this study are available from the corresponding author, NC, upon reasonable request.

---

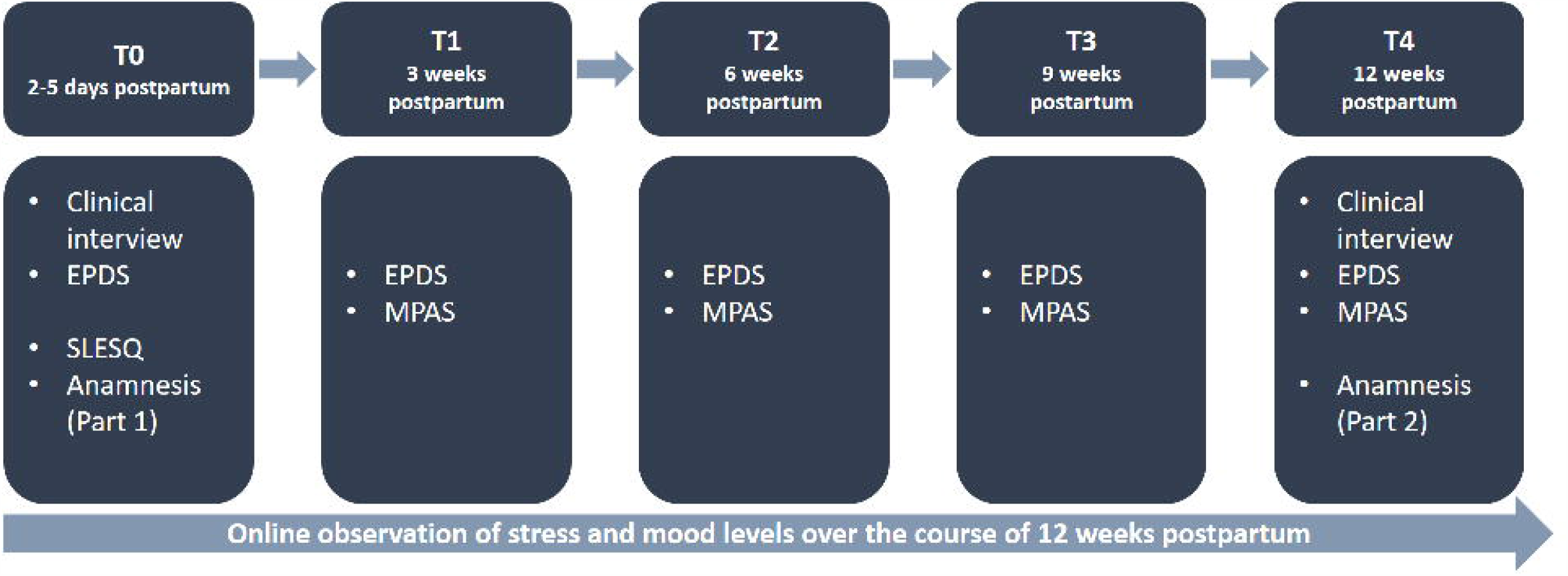

